# Clinical Profile of Vascular Behcet’s Disease Patients Over a 30-year Period: Insights from an Iranian Retrospective Cohort Study

**DOI:** 10.1101/2024.12.23.24319557

**Authors:** Soraya Shadmanfar, Kimia Jazi, Seyedeh Tahereh Faezi, Masoumeh Akhlaghi, Hamidreza Kelarestaghi, Zeynab Rastegar Moghadam, Fereydoun Davatchi, Seyed Mojtaba Alavi, Maryam Masoumi

## Abstract

Vascular manifestations are rare and cause morbidity and mortality in patients with Behçet’s disease (BD). Owing to the lack of evidence, we aimed to retrospectively evaluate the clinical features and associated risk factors of vascular BD. This retrospective cohort study was conducted at the Rheumatology Research Center, Tehran, Iran, from 1982 to 2018. The demographics, laboratory results, severity at diagnosis, diagnosis interval, and clinical manifestations divided into major events as well as sub-events at each visit were recorded. Of 57 patients, 43 (75.44%) were male, with an M:F ratio of 3.07. The mean age at onset was 32.00 ± 8.05 years. The three major clinical events repeated during the study period were mucocutaneous (41.30%), ocular (17%), and vascular (16.8%). DVT (33.3 %), SVT (21.84 %), and thrombosis of the large vein extremities (17.24 %) were the most common vascular events. The presence of HLA-B51 was found to have a statistically significant impact on the distinct number of events (B=-0.39, 95% CI: -0.74, -0.03, p=0.039). Severity at diagnosis (Moderate Vs. mild and Severe Vs. mild) were found to be statistically significant predictors of the logarithm of distinct number of events (B=-0.68, 95% CI: -1.13, -0.24, p=0.004; B=-0.51, 95% CI: -0.93, -0.10, p=0.020). The odds of developing vasculitis decreased faster in patients with a family history of OA than in those without a family history of OA. The other variables were not found to have a statistically significant impact on the logarithm of distinct number of events. A family history of OA, HLA- B51 positivity, and disease severity at diagnosis were associated with the occurrence of vascular BD. Further longitudinal and large-sample size studies should be conducted to evaluate the risk factors for vascular events.

## Introduction

Behçet’s disease (BD), a chronic multisystem complex vasculitis, is mainly characterized by recurrent oral and genital aphthous, cutaneous, ocular, and articular involvements [1]. Moreover, central and peripheral neurological, gastrointestinal, and cardiovascular manifestations have been reported [1, 2]. The worldwide prevalence of BD is 10.3/100 000 [3]. The estimated pooled prevalence was higher in countries around the SilkRoad, such as Turkey and Iran with 119.8 and 80 cases per 100 000 [3, 4].

Regarding a varied symptomatology, there are six phenotypes of BD including only mucocutaneous, predominant articular, ocular involvement (mostly associated with CNS manifestations and HLA-B51), dominant CNS, gastrointestinal, and vascular phenotype [5]. Most patients present with mucocutaneous ulcers [6, 7]. A cluster analysis in the Chinese population demonstrated five clusters, the most common of which were mucocutaneous, articular, gastrointestinal, ocular, and cardiovascular [8]. There are a considerable number of studies on the most frequent manifestation of the disease worldwide; however, vascular involvement, although decisive in prognosis and differential diagnosis, requires further investigation [7].

Vascular involvement is considered both a diagnostic factor and an important complication of Behcet’s disease. Studies have reported that 18-29% of patients with BD experience vascular involvement, which is one of the contributors to mortality among these patients [9, 10]. Histopathological assessment showed fibrous thickening of the vessels, aneurysm dilatation, thrombus formation, and inflammatory infiltration [11]. Vessels in all sizes and veins are affected; however, superficial and deep vein thrombosis (SVT, DVT) are the most prevalent [12]. DVT of the lower extremities is one of the earliest signs of vascular involvement in patients with BD [9]. Any arteries in the body can be involved, but they mostly affect the pulmonary artery and aorta [13]. Other rare arterial events, such as coronary artery disease, have also been reported [14]. Previous studies have shown a young male predominance of vascular involvement [12].

The quality of life of BD patients may be significantly reduced, which can impose a notable economic burden on both patients and nations [15]. The mortality rate associated with BD can increase under certain circumstances, particularly in young patients. The leading causes of death in these cases are large vessels and cardiac and neurological involvements [16]. The mortality rate of BD patients with vascular manifestations is 8% compared to 5-10% for all manifestations [9, 10]. Pulmonary artery aneurysm and Budd Chiari syndrome are two vascular involvements with the highest mortality (26-50% and 18-47% respectively) [17–19].

Regarding the poor prognosis of vascular BD and low prevalence of vascular BD, there is a paucity of studies evaluating the associated risk factors. In the current study, we conducted a comprehensive 30-year retrospective cohort analysis to identify the potential risk factors for vascular involvement in patients diagnosed with BD.

## Materials and methods

### 1. Study Design, settings, and patients

In this retrospective cohort study, patients with BD who had vascular involvement during their visits were included and assessed. This study was approved under the supervision of Qom University of Medical Sciences’ Research and Ethics (IR.MUQ.REC.1402.216), and conducted according to the Helsinki ethics criteria.

Data of patients with vascular BD from the Clinic of Rheumatology of Shariati Hospital between 1982 and 2018 were obtained retrospectively. All the data were gathered and analyzed anonymously; thus, the ethics committee waived the consent. Diagnoses of BD were established using the revised International Criteria for Behçet’s Disease (rICBD) by two different expert rheumatologists [20]. Patients with incomplete medical records and those with underlying diseases, such as malignancies, current infections, other autoimmune diseases, and endocrine diseases, were excluded from the study. Finally, a total of Fifty-seven patients were enrolled in the study.

### 2. Data extraction, and variables

Data extracted included the following information: demographics (such as sex, age at onset, smoking status, family history of BD, or oral aphthous (OA)) as well as the time interval between the first clinical presentation and the date of diagnosis, the presence of juvenile BD, and laboratory evaluations at diagnosis such as HLA-B5 and HLA-B5. The first two manifestations, including arthritis, mucocutaneous, constitutional, ocular, hematologic, neuropsychiatric, and vascular, that led to the BD diagnosis were also obtained. The severity of BD at diagnosis was evaluated using the Iranian Behçet’s disease dynamic activity measurement (IBDDAM) score at diagnosis [21]. The IBDDAM scores obtained for this study were based on active manifestations at the time of the first manifestation to the visit that led to BD diagnosis and did not affect the clinical decision-making and treatment of the patients. The scores were obtained in accordance with the examination profile in their clinical records at the time of diagnosis, as confirmed by two rheumatology experts. Further information is available regarding the scoring and investigation of disease activity using the IBDDM score [22].

During each visit during the observation period of 30 years, the clinical manifestations of the disease were recorded attentively and named events and sub-events. The events were classified into eight categories: skin, laboratory results, eye manifestations, vascular involvements, central nervous system (CNS) involvements, musculoskeletal manifestations, epididymitis, and renal involvement, etc. Specific clinical features were recorded as sub-events. Both the events and sub-events are listed in Table 5. To compare the associated risk factors and their effects on vascular involvement, patients were classified into large-, intermediate-, and small-vessel involvement groups. Large vessel involvement was defined as aneurysms of the CNS, abdominal aorta, thoracic aorta, and pulmonary artery, in addition to pseudoaneurysms of the subclavian artery, pulmonary artery embolism, thrombosis of the iliac artery, inferior vena cava (IVC), subclavian vein, and superior vena cava (SVC). Intermediate vessel involvement included DVT, pseudoaneurysm of the lower limbs, superficial vein thrombosis, and thrombosis of veins in the large extremities. Small vessel involvements included iliac, jugular, upper limb, visceral vein thrombosis, and aneurysms of the renal artery.

### 3. Statistical Analysis

Descriptive statistics were utilized, presenting symmetric and asymmetric numeric variables as mean ± standard deviation (SD) and median (interquartile range [IQR]). Categorical variables were presented as frequencies (percentages). Descriptive statistics of events were reported, considering repeated cases for each participant across time. The percentage of first manifestations was computed as shown in Fig S1. Similarly, the yearly percentage of each event was determined, as shown in Fig S2. An evaluation of the relationship between the first manifestations and variables, including family history of oral aphthous and Bechet’s disease (BD), was performed using the Fisher-Freeman-Halton Exact test. The presentation of the time intervals between the diagnosis date and the first manifestation and between the diagnosis date and the first event was organized according to the first manifestation factors. In addition, the frequency of each sub-event was documented for the main events. To assess the impact of variables on vascular events, as they evolve over time and vary among participants, a generalized linear mixed-effects model (GLMM) was employed. The effect of each variable on the number of distinct events was examined through a simple regression analysis involving a logarithmic transformation to normalize the response. Statistical analysis was conducted using R (version 4.3.1), with p-values less than 0.05 being considered as indicative of statistical significance.

## Results

### 1. Descriptive Statistics

As shown in Table 1, Of the 57 patients, 43 (75.44%) patients were male, and 14 (24.56%) were female. The mean (± SD) age of patients was 32.00 ± 8.05 years. The majority of patients were Turk (43.86%), followed by Fars (24.56%) and Lor (5.26%). Only one patient had juvenile BD (1.75%). A substantial proportion of patients had no family history of oral aphthous ulcers (71.93%), whereas 28.07% reported a positive family history. The majority had no family history of Behçet’s disease (98.25%) and a smaller percentage reported such a history (1.75%). All patients were non-smokers. HLA-B5 was negative in 27 patients (47.37%) and positive in 30 (52.63%). HLA-B51 was negative in 33 patients (57.89%) and positive in 24 (42.11%). The severity of diagnosis was categorized as mild (38.60%), moderate (24.56%), or severe (31.58%). The distribution of disease manifestations by year and participant ID revealed that skin manifestations were the most common (41.13%), followed by eye (7.00%) and vascular (16.80%) events.

**Table 1.** Descriptive characteristics of patients with Behcet’s disease.

| Variable | Levels | Frequency (percentage) |
| --- | --- | --- |
| Age at onset | Mean ± SD | 32.00 ± 8.05 |
| Sex | Female | 14 (24.56) |
|  | Male | 43 (75.44) |
| Family history of Oral Aphthous | No | 41 (71.93) |
|  | Yes | 16 (28.07) |
| Family history of BD | No | 56 (98.25) |
|  | Yes | 1 (1.75) |
| Juvenile BD | No | 56 (98.25) |
|  | Yes | 1 (1.75) |
| Smoking | No | 57 (100.00) |
|  | Yes | 0 (0.00) |
| HLAB5 | No | 27 (47.37) |
|  | Yes | 30 (52.63) |
| HLAB51 | No | 33 (57.89) |
|  | Yes | 24 (42.11) |
| Severity at diagnosis | Mild | 22 (38.60) |
|  | Moderate | 14 (24.56) |
|  | Severe | 18 (31.58) |
| Event* | Skin | 204 (41.30) |
|  | Musculoskeletal | 20 (4.05) |
|  | Vascular | 83 (16.80) |
|  | Eye | 84 (17.00) |
|  | Varicocele | 3 (0.61) |
|  | CNS | 10 (2.02) |
|  | Surgery | 1 (0.20) |
|  | Infection | 4 (0.81) |
|  | GI | 1 (0.20) |
|  | Epididymitis | 4 (0.81) |
|  | Renal | 6 (1.21) |
|  | Soft tissue abscess | 1 (0.20) |
|  | HTN | 3 (0.61) |
|  | Lung | 4 (0.81) |
|  | Heart | 1 (0.20) |

Note:
Categorical variables are presented as frequencies (percentages). Numeric variables are summarized as mean ± SD. The distinct participants are included in the above table. Abbreviation: SD, standard deviation (SD). \*All participants with repeated events are presented.

### 2. Temporal Distribution of First Manifestations in Behcet’s Disease

Table 2 provides a comprehensive overview of the initial manifestations of Behçet’s disease, categorized into the first 1-manifestation and first 2-manifestation symptoms documented across multiple years. Notably, the highest percentages in the first 1-manifestation and first 2- manifestation categories were observed for mucocutaneous symptoms, accounting for 82.46% and 63.16%, respectively. Vascular manifestations showed a notable increase from 5.26% in the first 1-manifestation to 14.04% in the first 2-manifestation cases. Similarly, Ocular symptoms increased from 3.51% in the first 1-manifestation to 7.02% in the first 2-manifestation cases. Arthritis was present as the initial manifestation in 1.75% of the first 1-manifestation cases but was entirely absent in the first 2-manifestation cases (0.00%). It is noteworthy that cases with missing data were significant in both groups, constituting 7.02% in the first 1-manifestation and 15.79% in the first 2-manifestation cases. This table and Fig S1 also represent the frequency of the initial manifestations of Bechet’s disease across multiple years.

**Table 2.**
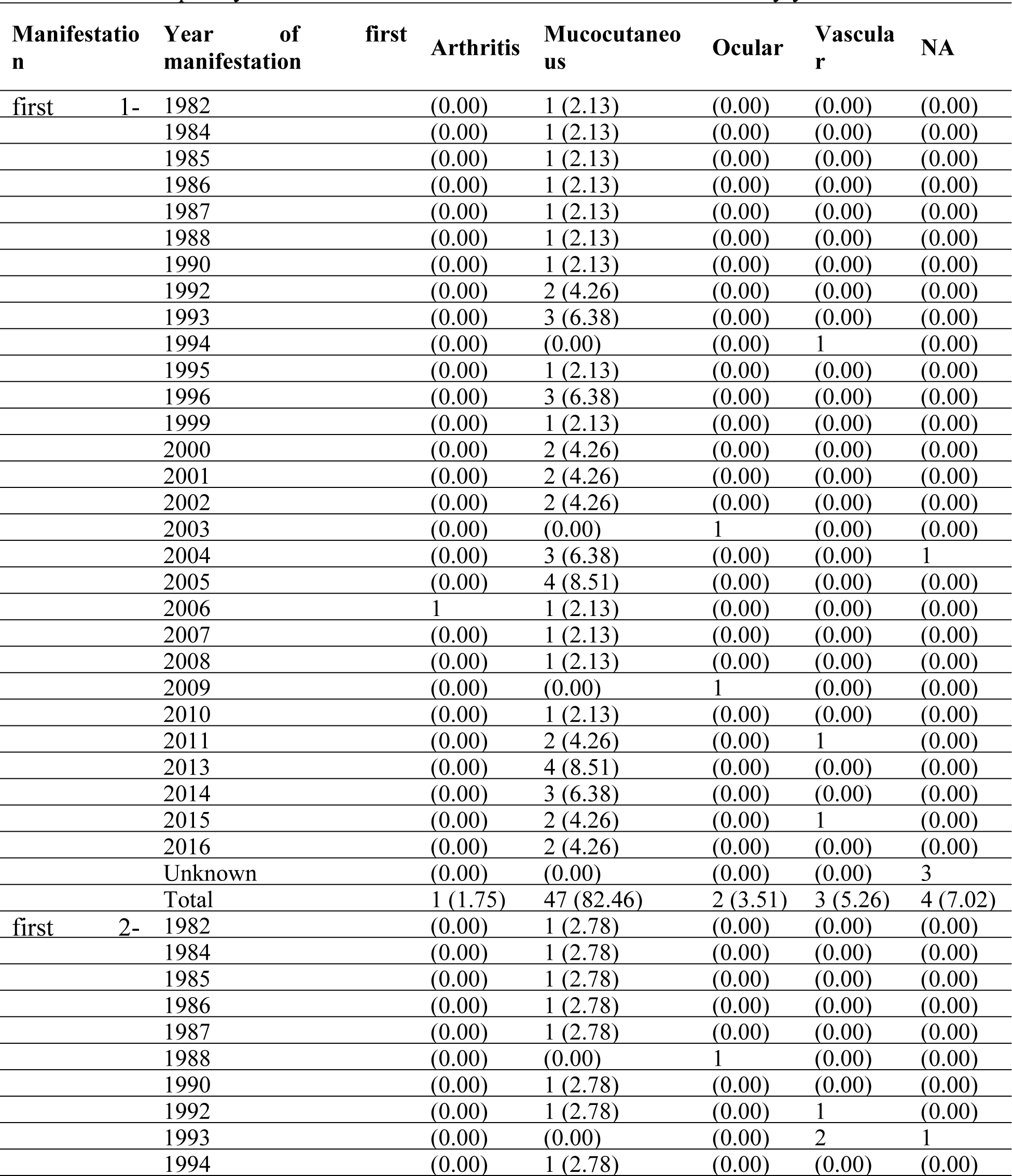
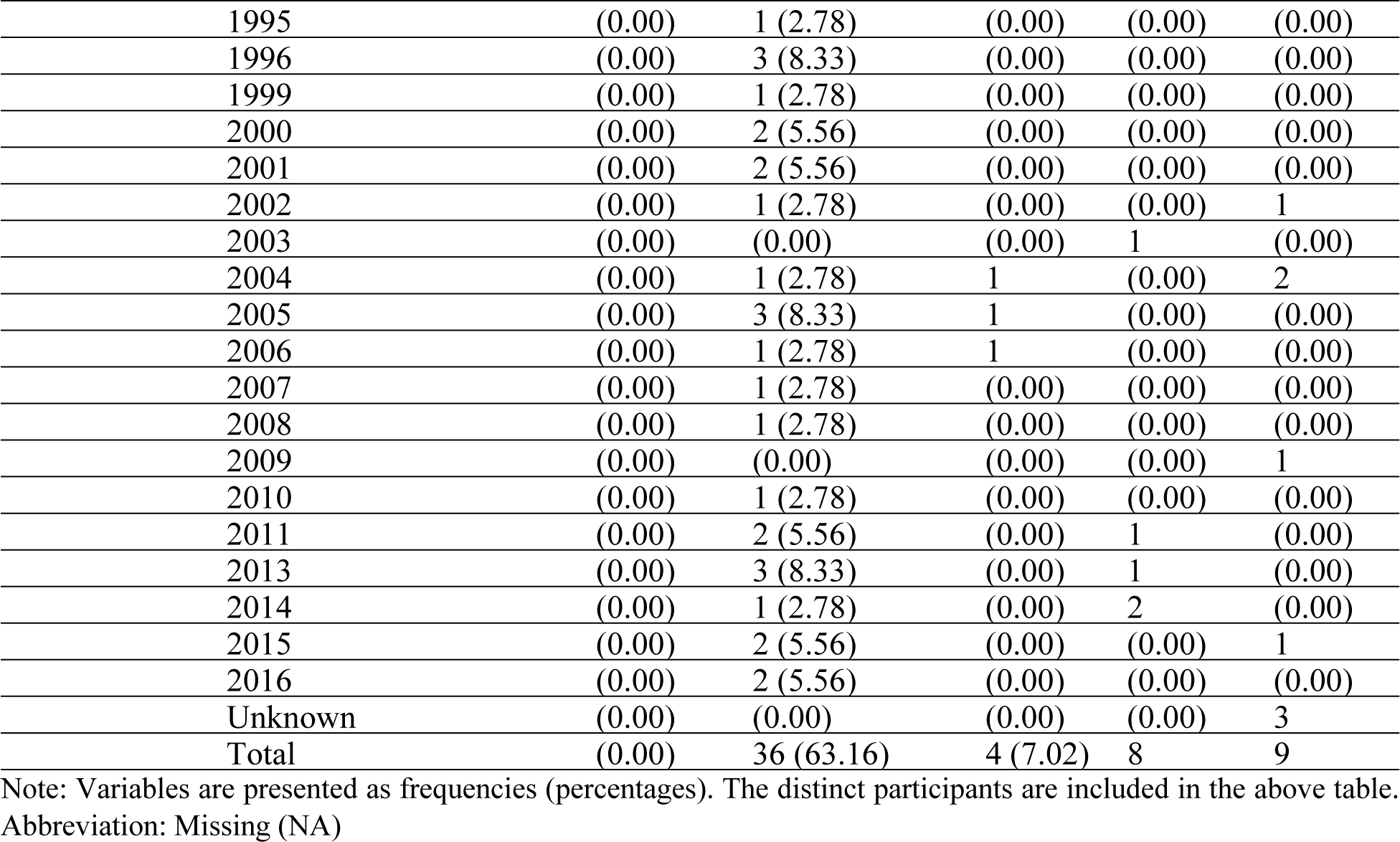
The frequency of first 1-manifestation and first 2-manifestation by year.

| Manifestation | Year of first manifestation | Arthritis | Mucocutaneous | Ocular | Vascular | NA |
| --- | --- | --- | --- | --- | --- | --- |
| first 1- | 1982 | (0.00) | 1 (2.13) | (0.00) | (0.00) | (0.00) |
|  | 1984 | (0.00) | 1 (2.13) | (0.00) | (0.00) | (0.00) |
|  | 1985 | (0.00) | 1 (2.13) | (0.00) | (0.00) | (0.00) |
|  | 1986 | (0.00) | 1 (2.13) | (0.00) | (0.00) | (0.00) |
|  | 1987 | (0.00) | 1 (2.13) | (0.00) | (0.00) | (0.00) |
|  | 1988 | (0.00) | 1 (2.13) | (0.00) | (0.00) | (0.00) |
|  | 1990 | (0.00) | 1 (2.13) | (0.00) | (0.00) | (0.00) |
|  | 1992 | (0.00) | 2 (4.26) | (0.00) | (0.00) | (0.00) |
|  | 1993 | (0.00) | 3 (6.38) | (0.00) | (0.00) | (0.00) |
|  | 1994 | (0.00) | (0.00) | (0.00) | 1 | (0.00) |
|  | 1995 | (0.00) | 1 (2.13) | (0.00) | (0.00) | (0.00) |
|  | 1996 | (0.00) | 3 (6.38) | (0.00) | (0.00) | (0.00) |
|  | 1999 | (0.00) | 1 (2.13) | (0.00) | (0.00) | (0.00) |
|  | 2000 | (0.00) | 2 (4.26) | (0.00) | (0.00) | (0.00) |
|  | 2001 | (0.00) | 2 (4.26) | (0.00) | (0.00) | (0.00) |
|  | 2002 | (0.00) | 2 (4.26) | (0.00) | (0.00) | (0.00) |
|  | 2003 | (0.00) | (0.00) | 1 | (0.00) | (0.00) |
|  | 2004 | (0.00) | 3 (6.38) | (0.00) | (0.00) | 1 |
|  | 2005 | (0.00) | 4 (8.51) | (0.00) | (0.00) | (0.00) |
|  | 2006 | 1 | 1 (2.13) | (0.00) | (0.00) | (0.00) |
|  | 2007 | (0.00) | 1 (2.13) | (0.00) | (0.00) | (0.00) |
|  | 2008 | (0.00) | 1 (2.13) | (0.00) | (0.00) | (0.00) |
|  | 2009 | (0.00) | (0.00) | 1 | (0.00) | (0.00) |
|  | 2010 | (0.00) | 1 (2.13) | (0.00) | (0.00) | (0.00) |
|  | 2011 | (0.00) | 2 (4.26) | (0.00) | 1 | (0.00) |
|  | 2013 | (0.00) | 4 (8.51) | (0.00) | (0.00) | (0.00) |
|  | 2014 | (0.00) | 3 (6.38) | (0.00) | (0.00) | (0.00) |
|  | 2015 | (0.00) | 2 (4.26) | (0.00) | 1 | (0.00) |
|  | 2016 | (0.00) | 2 (4.26) | (0.00) | (0.00) | (0.00) |
|  | Unknown | (0.00) | (0.00) | (0.00) | (0.00) | 3 |
|  | Total | 1 (1.75) | 47 (82.46) | 2 (3.51) | 3 (5.26) | 4 (7.02) |
| first 2- | 1982 | (0.00) | 1 (2.78) | (0.00) | (0.00) | (0.00) |
|  | 1984 | (0.00) | 1 (2.78) | (0.00) | (0.00) | (0.00) |
|  | 1985 | (0.00) | 1 (2.78) | (0.00) | (0.00) | (0.00) |
|  | 1986 | (0.00) | 1 (2.78) | (0.00) | (0.00) | (0.00) |
|  | 1987 | (0.00) | 1 (2.78) | (0.00) | (0.00) | (0.00) |
|  | 1988 | (0.00) | (0.00) | 1 | (0.00) | (0.00) |
|  | 1990 | (0.00) | 1 (2.78) | (0.00) | (0.00) | (0.00) |
|  | 1992 | (0.00) | 1 (2.78) | (0.00) | 1 | (0.00) |
|  | 1993 | (0.00) | (0.00) | (0.00) | 2 | 1 |
|  | 1994 | (0.00) | 1 (2.78) | (0.00) | (0.00) | (0.00) |

|  |  |  |  |  |  |
| --- | --- | --- | --- | --- | --- |
| 1995 | (0.00) | 1 (2.78) | (0.00) | (0.00) | (0.00) |
| 1996 | (0.00) | 3 (8.33) | (0.00) | (0.00) | (0.00) |
| 1999 | (0.00) | 1 (2.78) | (0.00) | (0.00) | (0.00) |
| 2000 | (0.00) | 2 (5.56) | (0.00) | (0.00) | (0.00) |
| 2001 | (0.00) | 2 (5.56) | (0.00) | (0.00) | (0.00) |
| 2002 | (0.00) | 1 (2.78) | (0.00) | (0.00) | 1 |
| 2003 | (0.00) | (0.00) | (0.00) | 1 | (0.00) |
| 2004 | (0.00) | 1 (2.78) | 1 | (0.00) | 2 |
| 2005 | (0.00) | 3 (8.33) | 1 | (0.00) | (0.00) |
| 2006 | (0.00) | 1 (2.78) | 1 | (0.00) | (0.00) |
| 2007 | (0.00) | 1 (2.78) | (0.00) | (0.00) | (0.00) |
| 2008 | (0.00) | 1 (2.78) | (0.00) | (0.00) | (0.00) |
| 2009 | (0.00) | (0.00) | (0.00) | (0.00) | 1 |
| 2010 | (0.00) | 1 (2.78) | (0.00) | (0.00) | (0.00) |
| 2011 | (0.00) | 2 (5.56) | (0.00) | 1 | (0.00) |
| 2013 | (0.00) | 3 (8.33) | (0.00) | 1 | (0.00) |
| 2014 | (0.00) | 1 (2.78) | (0.00) | 2 | (0.00) |
| 2015 | (0.00) | 2 (5.56) | (0.00) | (0.00) | 1 |
| 2016 | (0.00) | 2 (5.56) | (0.00) | (0.00) | (0.00) |
| Unknown | (0.00) | (0.00) | (0.00) | (0.00) | 3 |
| Total | (0.00) | 36 (63.16) | 4 (7.02) | 8 | 9 |

Note: Variables are presented as frequencies (percentages). The distinct participants are included in the above table. Abbreviation: Missing (NA)

#### **a.** The Association Between First Manifestation and Family History of OA and BD

Table 3 presents the frequency and percentage of family history of oral aphthous (OA) and Behçet’s disease (BD) by the first manifestation of the disease. This table also provides an analysis of the relationship between family history and the various levels of initial manifestation. The findings revealed that there was no statistically significant association between the occurrence of oral aphthous (OA) or Behcet’s disease (BD) and the levels of manifestation, irrespective of whether it is the first manifestation (1-Manifestation) or the second manifestation (2- Manifestation).

**Table 3.**
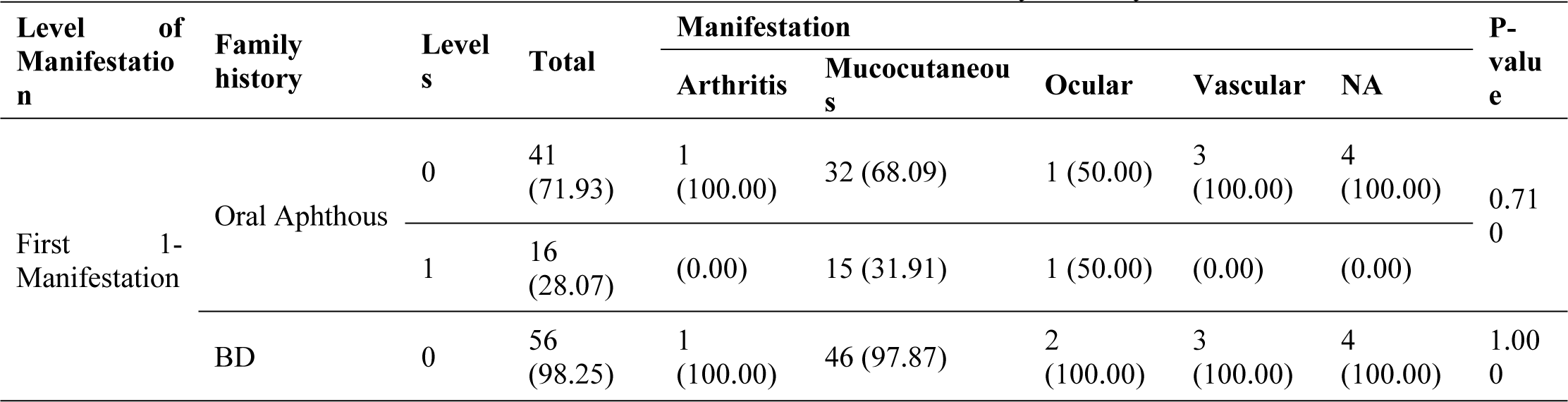
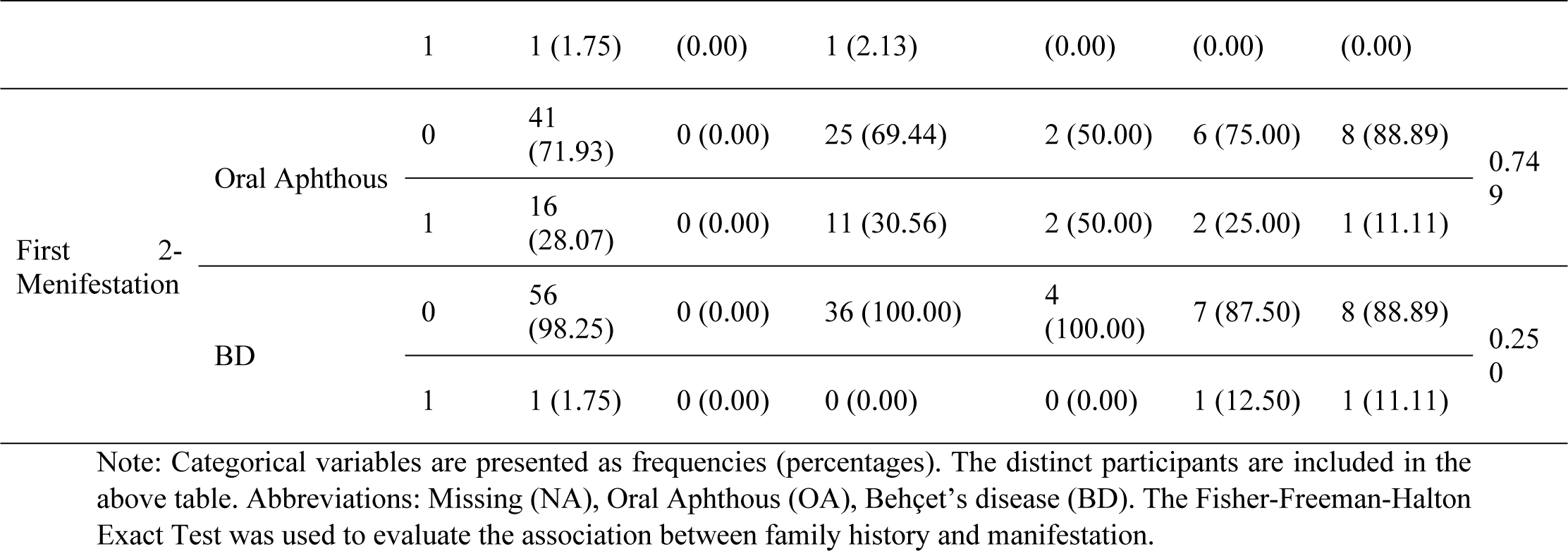
The Association Between First Manifestation and Family History of OA and BD.

#### **b.** Temporal Trends in Medical Event Prevalence

Fig S2 (and Table S2 in the supplementary file) present a concise overview of the annual percentage distribution of medical events across various organ systems and categories from 1988 to 2018. The prevalence of these events varies widely over the years, with notable trends including consistent occurrences of “Skin” and “Musculoskeletal” conditions, sporadic appearances of “Surgery” and “Infection,” and fluctuations in the “Vascular” and “Eye” categories. “Hypertension (HTN)” exhibits a significant increase starting from 2000, while “Renal” conditions peak at 100.00% in 1991. The year 2018 was exceptional, with a singular event in the “Vascular” category at 100.00%.

#### **c.** Temporal Variability in Disease Progression Across Different First Manifestation Factors

Table 4 presents the descriptive statistics of the time interval between the first manifestation and diagnosis date, categorized by the first manifestation factors. The median time interval for arthritis was 1 (IQR: 1, 1), for mucocutaneous was 2 (IQR: 1, 6.75), for ocular was 2.5 (IQR: 1.25, 3.75), and for vascular was 3 (IQR: 2.5, 3.5). The median time interval for all manifestations combined was two (IQR: 1, 6). In the context of the diagnosis date and the first event, Mucocutaneous patients displayed a median time interval of 0.00 (IQR: 0.00, 0.14). Similarly, Ocular patients had a median of 1.95 (IQR: 0.98, 2.93). Vascular patients exhibited the longest median time interval of 5.45 (IQR: 2.75, 8.15). Overall, the total mean time interval for all patients across the first event factors was 0.98 ± 2.45, reflecting variations in the timing of events across different first manifestation groups.

**Table 4.** Time Intervals Between Diagnosis Date and First Manifestation, and Diagnosis Date and First Event Across First Manifestation Factors manifestation.

| Group | Variable | Mean ± SD | Median (IQR) |
| --- | --- | --- | --- |
| diagnosis date to first manifestation | Arthritis | 1.00 ± 0.00 | 1 (1, 1) |
| | Mucocutaneous | 5.11 ± 6.88 | 2 (1, 6.75) |
| | Ocular | 2.50 ± 3.54 | 2.5 (1.25, 3.75) |
| | Vascular | 3.00 ± 1.00 | 3 (2.5, 3.5) |
| | **Total** | 4.85 ± 6.49 | 2 (1, 6) |
| diagnosis data and first event | Arthritis | ----- | ----- |
| | Mucocutaneous | 0.78 ± 2.07 | 0.00 (0.00, 0.14) |
| | Ocular | 1.95 ± 2.76 | 1.95 (0.98, 2.93) |
| | Vascular | 5.45 ± 7.63 | 5.45 (2.75, 8.15) |
| | **Total** | 0.98 ± 2.45 | 0.00 (0.00, 0.15) |

Abbreviations: standard deviation (SD), interquartile range (IQR), standard deviation

#### **d.** Distribution of Sub-Events within Main Events: Frequency and Percentage Analysis

Table 5 provides a comprehensive overview of the frequency and percentage distributions of the sub-events within the various main events. Notably, the majority of skin events were predominantly associated with sub-events, such as Oral Aphthous (58.33%), Erythema Nodosum (24.51%), and Genital Aphthous (9.31%). Vascular main events had the highest frequencies in DVT (33.33%), superficial vein thrombosis (21.84%), and thrombosis/vein/large extremities (17.24%). Sub-events with the highest percentages of occurrence included retinal/vasculitis (45.24%), uveitis/posterior (26.19%), and retinal/pylephlebitis (11.90%). The main musculoskeletal event was predominantly characterized by Mono Arthritis, which accounted for 50.00% of its occurrences.

**Table 5.**
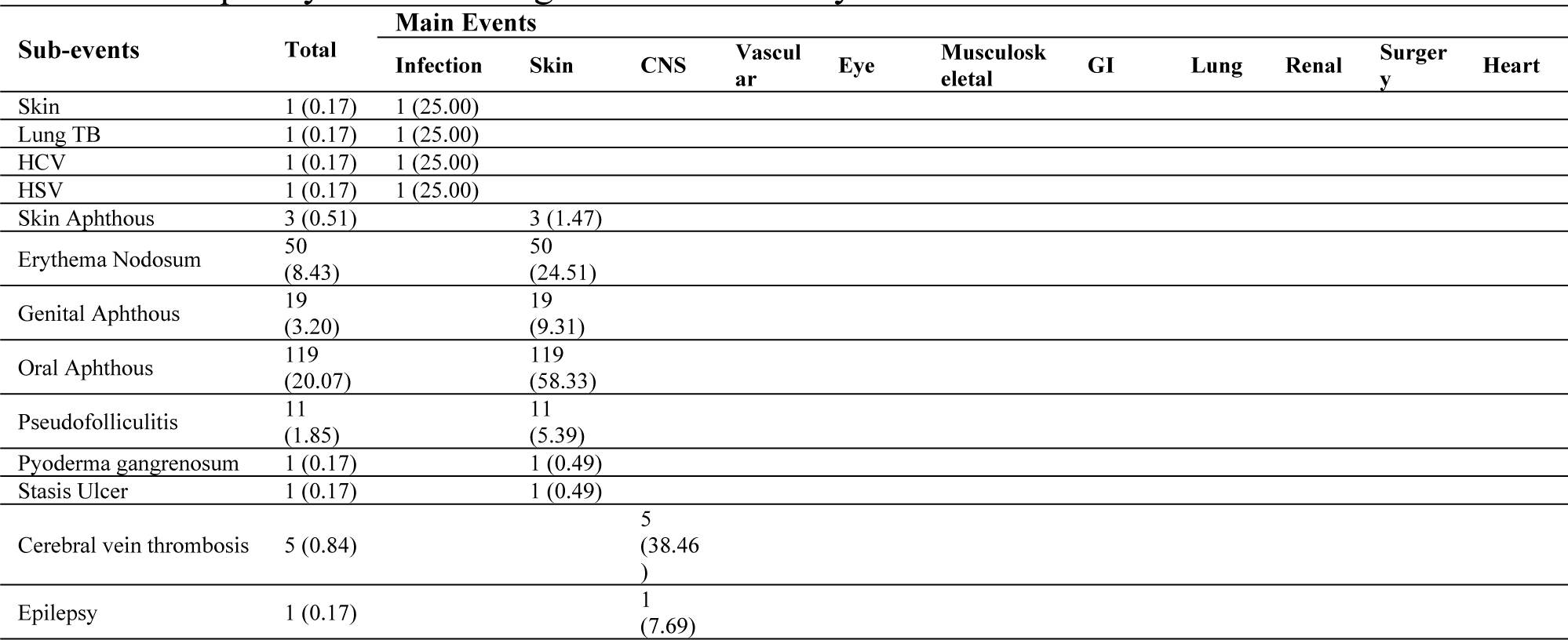
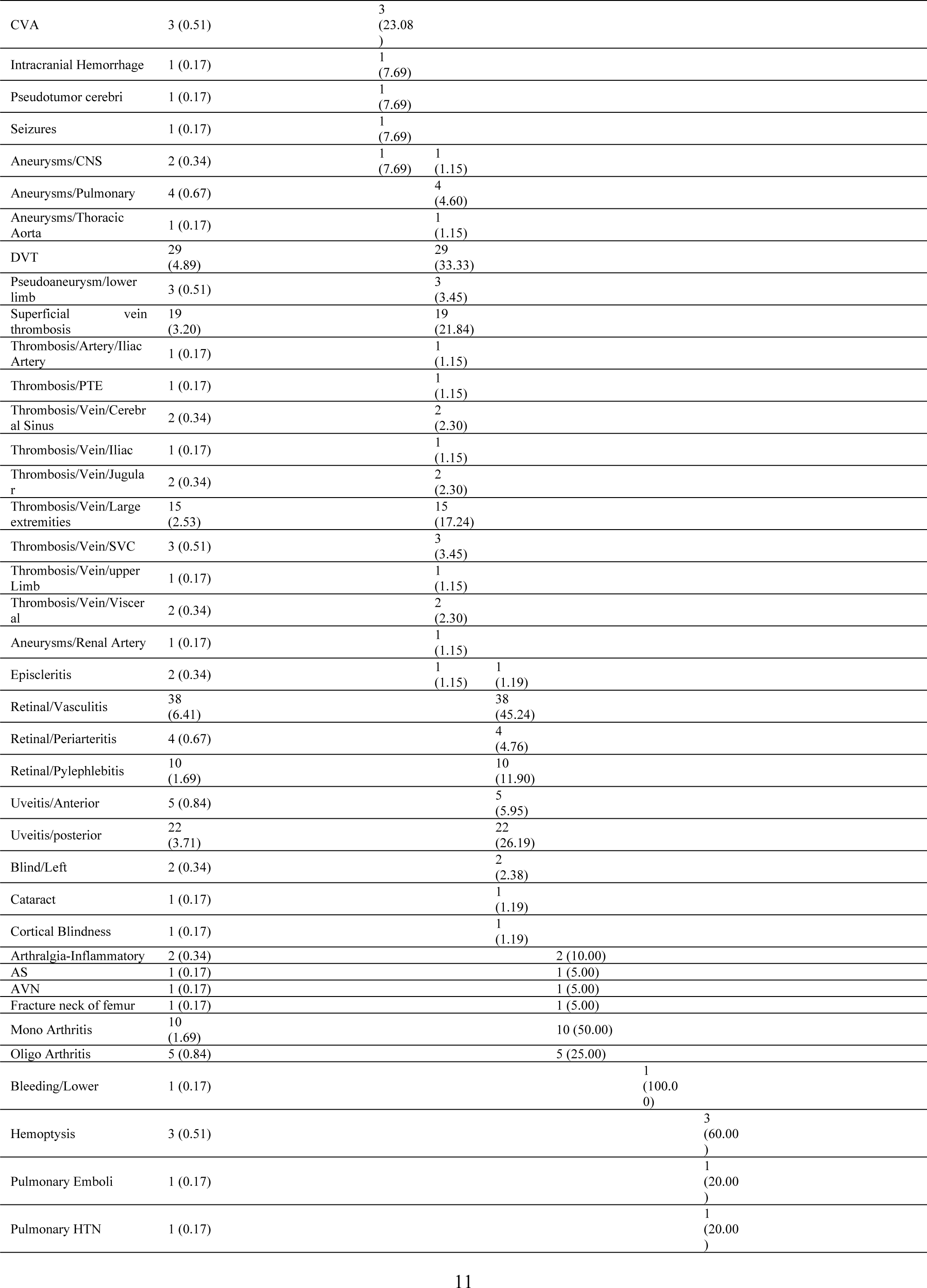
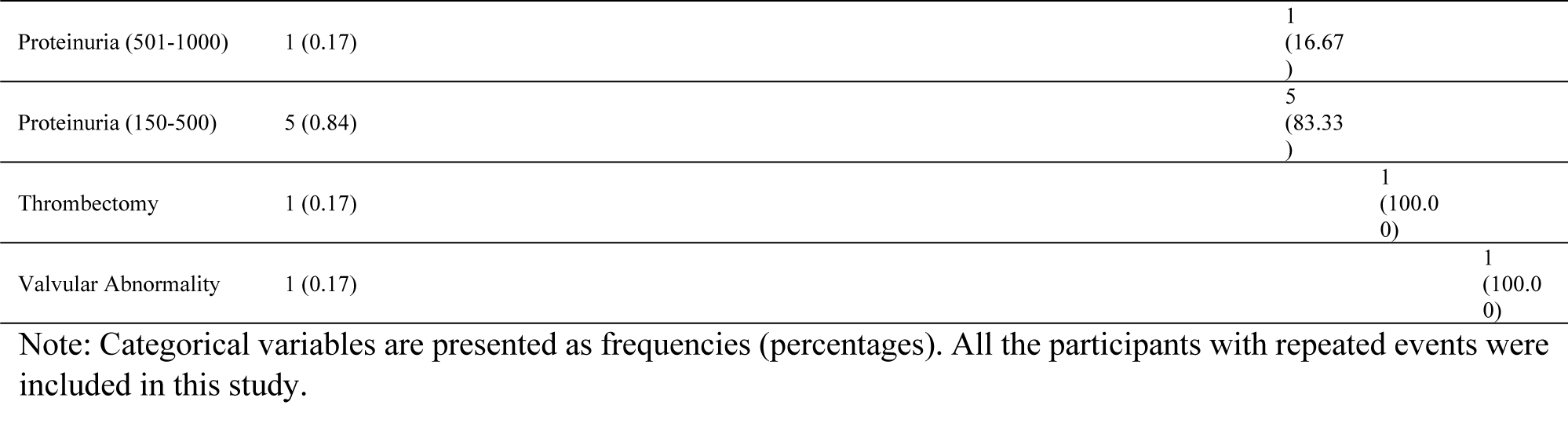
Frequency and Percentage of Sub-Events by Main Events.

#### **e.** Impact of Variables on Events

In Table 6.1, the impact of various variables on vascular events was evaluated using a generalized linear mixed-effects model (GLMM) that considered the change in this event over time and by participant ID. The variables evaluated were age at onset, event year, sex, juvenile BD, HLAB5, HLAB51, family history (FH) of OA, FH of BD, and severity at diagnosis. P-values less than 0.05 were considered statistically significant. The results showed that none of the variables had a statistically significant impact on vascular events, except for the FH of OA interaction with event year (OR = 0.90, 95% CI = 0.81-0.99, p = 0.030, respectively). This means that the odds of developing vascular disease declined more rapidly among patients with family history of OA than to those without family history of OA. In addition, Table S3 (Supplementary File) shows that age at onset, juvenile BD, HLAB51, FH of BD, and severity at diagnosis did not have a statistically significant impact on vascular events. However, the odds of a vascular event occurring were 75% higher in men than in women (OR, 1.75; 95% CI, 1.00, 3.07; p=0.049).

**Table 6.**
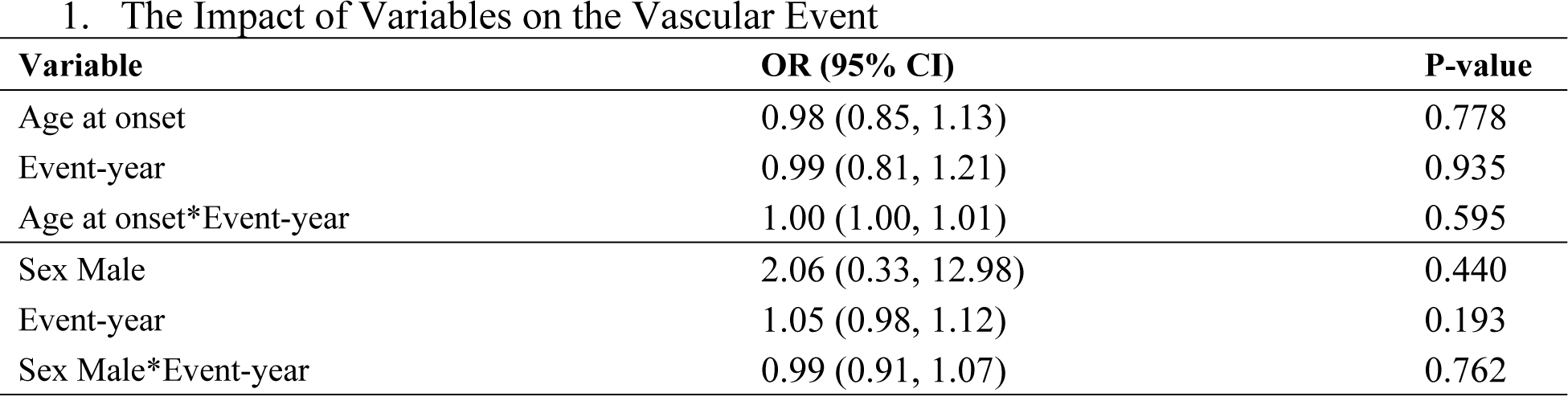
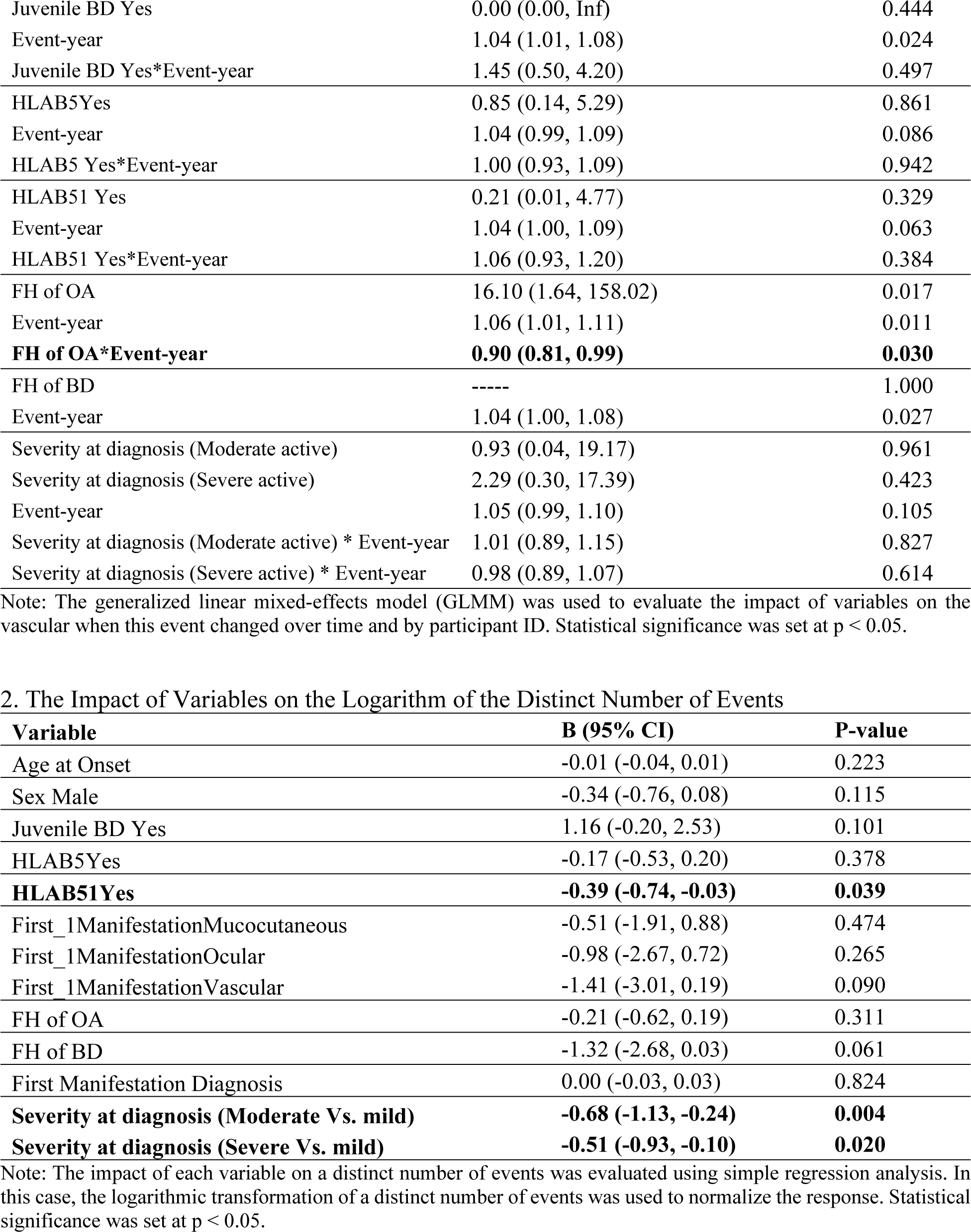
The Impact of Variables on the Vascular Event.

Table 6.2 also shows the impact of the various variables on the logarithm of the number of distinct events. Among the variables evaluated, the presence of HLAB51 was found to have a statistically significant impact on the number of events (B=-0.39, 95% CI: -0.74, -0.03, p=0.039). Also, Severity at diagnosis (Moderate Vs. mild) and Severity at diagnosis (Severe Vs. mild) were found to be statistically significant predictors of the logarithm of distinct number of events (B=-0.68, 95% CI: -1.13, -0.24, p=0.004; B=-0.51, 95% CI: -0.93, -0.10, p=0.020). The other variables were not found to have a statistically significant impact on the logarithm of distinct number of events.

## Discussion

Compared with other inflammatory vascular conditions, BD exhibits unique features. It can affect any type of blood vessel and is more likely to form thrombosis. It predominantly involves veins rather than arteries [12]. Vascular events occur in up to 50% of patients [12]. There is not enough epidemiological data on vascular BD in the literature. This is the first retrospective cohort analysis of the clinical presentations of patients with vascular BD and associated risk factors.

The mean male-to-female ratio was 3.07. Sex differences among patients with BD depend on their country and ethnic origin [23]. Male patients were more dominant among Turkish population in comparison to Americans (27% vs. 87%) [24]. The male-to-female ratios were 0.63 in Korea [25], 0.98 in Japan [26], and 1.2 reported by Rheumatology Research Center report of Iran [27]. The high male-to-female ratio in developing and under-developed countries such as North Africa, Turkey, and Iran could be due to the reluctance of women with genital ulcers to seek medical care [23, 28]. Hormonal differences may also explain why vascular events are more common in men [29]. Torgutalp et al. evaluated 460 patients with BD with vascular involvement and discovered that male sex and SVTs were major risk factors for vascular events [30]. Consistently, Alkaabi et al. reported that male sex and disease duration were significantly associated with future vascular events [31]. Atalar et al. also suggested that male and older patients with BD are more vulnerable to vascular involvement [32]. Although 75% of our patients with vascular events were men, we did not find that male sex was a risk factor for developing vascular involvement. These disparities might be due to the small sample size and the population-specific features of the disease.

Our findings showed that the most common initial presentation was mucocutaneous, representing 82.46% of the initial manifestations of the disease. Similarly, another study on BD patients indicated that oral ulcers, genital ulcers, eye involvement, and other skin involvements were the most frequent first presentations before the diagnosis of complete BD [33]. Balt et al. reported that the majority of their patients represented with oral aphthous (61.5%) as the initial symptom. They did not report any cases with initial symptom of vascular involvement [34]. Khabazi et al. demonstrated that 83.1% of their patients had OA as the initial symptom, with only 0.6% presenting with vascular involvement [35]. Ryu et al. identified vascular involvement as the primary symptom in 3.1% of Korean patients [36]. Alakkas et al. reported a 22-year-old Arabian man presented with pulmonary artery thrombosis as the first manifestation of BD [37]. Interestingly, 5.26% and 14.04 % of our patients presented with vascular involvement as their two first disease manifestation, respectively. Vascular BD are more prevalent among young males, it has been reported to manifest their first vascular event within five years of disease onset, of whom 10.8% present with vascular events before achieving the diagnostic criteria [9, 38, 39].

In an analysis of 421 Japanese patients with BD, vascular events occurred after an average of 2.2±8.4 years following BD diagnosis. Venous involvement tended to occur in a shorter period compared to arterial involvement, although no significant difference was observed (median: 0.3 vs 1.9, respectively) [40]. Similarly, the current results demonstrated that the median interval between diagnosis and occurrence of vascular event was 5.45 years. Moreover, the median time interval from the first manifestation to the diagnosis of BD was 3 years among Iranian vascular BD patients. However, Ideguchi et al. through a study on 412 patients with BD in 16 years, reported that time from the first symptom to diagnosis was 8.6 ± 10.1 years [33]. Based on our findings, vascular events often occur as late-onset complications and are associated with higher mortality and morbidity rates. Patients who develop vascular involvement also have a shorter diagnostic time interval than those with no vascular signs. To date, vascular lesions are not included in International Study Group for Behçet’s disease (ISG) diagnostic criteria, further, The International Criteria for Behçet’s Disease (ICBD) weight half of other manifestations [41, 42]. We suggest that further longitudinal large-scale studies should focus on adding vascular events with a population-based proper weight in the BD diagnostic criteria to achieve early diagnosis, early treatment, and consequently, decrease disease-associated morbidity.

The three major clinical events repeated during our study period were mucocutaneous (41.30%), ocular (17%), and vascular (16.8%). Vascular lesions accounted for 14% of 2,319 patients in Turkey. Moreover, SVT, DVT, and arterial lesions were the most commonly reported [43]. Among the Chinese BD population, 13% of 796 patients showed vascular manifestations, of which arterial lesions, venous lesions, and both were the most frequent [38]. Large vessel involvement has also been reported in one-third of the patients with BD [44]. Although there have been multiple reports of pulmonary artery aneurysm in Western countries, it is uncommon among Japanese [37, 45]. Tascilar et al. demonstrated that most vascular Turkish BD patients (87%) had DVT [9]. Recently, Atalar et al. reported that approximately 70% of vascular involvements in BD patients were DVT, with 12.9% involving the pulmonary arteries [32]. They also found that younger patients were at a higher risk of DVT, whereas older patients were more likely to develop pulmonary artery involvement [32]. According to our findings, DVT (33.3 %), SVT (21.84 %), and thrombosis of the large vein extremities were observed in Iranian patients with vascular BD (17.24%). Of all the sub-events, DVT accounted for (4.89%) subsequent to oral aphthous (20.07%), erythema nodosum (8.43%), and retinal vasculitis (6.41%).

HLA-B51 was positive in 42.11% of the patients. In contrast to previous studies, we found that HLA-B51 positivity was negatively associated with the occurrence of more events in vascular BD patients. However, it was not significantly associated with vascular involvements in particular. In addition to the male sex, the presence of HLA-B51 has been reported to be associated with any vascular events but major events [30]. Pamukcu et al. revealed that HLA B51 was correlated with more papulopustular lesions and vascular, ocular, and neurological involvements in BD patients [39]. Nonetheless, Erdem Sultanoğlu et al. reported that thrombophlebitis was more common in HLA-B51 positive patients, but it was not considered a risk factor [46]. Consistent with our results, recently, Leccese et al., subtyping HLA-B51 among 241 Italian BD patients, underlined that the B51:01 and B51:08 subtypes were associated with ocular involvement and had no significant association with vascular involvement [47]. There is still a lack of evidence that what subtype of HLA-B51 is associated with each manifestation. Moreover, disparities in the results, the small size of our study, and the scarce occurrence of vascular BD highlight the importance of further large-scale longitudinal observations.

28.07% of our patients had a family history of OA. Although recurrent OA is a well-known manifestation of BD, surprisingly, vascular events are less likely to occur in these patients. Regarding limitations, the exact pathogenesis remains unknown, and further investigations should be conducted to confirm this result.

The present study had some limitations. First, it was conducted at a single center, which may limit the generalizability of the results. Single-center studies may not fully capture the diversity of clinical presentations and outcomes across different geographic and ethnic populations. Further multicenter studies are needed to validate these findings in diverse patient populations. Second, retrospective studies rely on existing records, which may not include all relevant data or may suffer from inconsistencies in how the data were recorded. Further prospective studies are needed to accurately determine the onset and progression of vascular BD and its associated risk factors. Third, the sample size might have resulted in a low statistical power to detect other associations and outcomes. Larger cohorts are necessary to confirm these risk factors and uncover additional insights. Fourth, our study might have some degree of selection bias owing to the exclusion of patients with incomplete medical records and underlying diseases. In summary, further cohort studies with a larger study population and multiple centers are needed to accurately identify factors related to vascular involvement in patients with BD. Geographic, ethnic, socioeconomic, and cultural differences, along with historical and genetic differences, should be evaluated.

## Conclusion

DVT and SVT are the most common vascular manifestations of BD in an Iranian population. HLA- B51 positivity and disease severity at the time of diagnosis are associated with a number of vascular events. The odds of developing vasculitis decreased faster in patients with a family history of OA than in those without a family history of OA. Further longitudinal and large-sample size studies should be conducted to evaluate the risk factors for vascular events.

## Acknowledgements

Special thanks to the Tehran Rheumatology Research Center, Clinical Research of Development Unit of Shahid Beheshti Hospital of Qom, as well as to all the patients who altruistically made this study possible.

## Ethical Standards

This study was approved under the supervision of Qom University of Medical Sciences’ Research and Ethics (IR.MUQ.REC.1402.216), and conducted according to the Helsinki ethics criteria. All the data were gathered and analyzed anonymously; thus, the ethics committee waived the consent.

## Data Availability

The data is available on the reasonable request from corresponding author.

## Author Contributions

Conceptualization: Soraya Shadmanfar, Kimia Jazi, Maryam Masoumi; Methodology: Maryam Masoumi, Massoumeh Akhlaghi, Hamidreza Kelarestaghi; Formal analysis and investigation: Kimia Jazi, Seyed Mojtaba Alavi, Seyedeh Tahereh Faezi; Writing - original draft preparation: Soraya Shadmanfar, Seyedeh Tahereh Faezi, Maassoumeh Akhlaghi, Hamidreza Kelarestaghi, Zeynab Rastegar Moghadam; Writing - review and editing: Kimia Jazi, Seyed Mojtaba Alavi, Fereydoun Davatchi; Resources: [full name], …; Supervision: Fereydoun Davatchi, Maryam Masoumi

## Supporting Information

**S1 Table.** The frequency (percentage) of disease severity by year of diagnosis Note: Categorical variables are presented as frequencies (percentages). The distinct participants are included in the above table. Abbreviation: Missing (NA).

**S2 Table.** Yearly Event Percentage Distribution Note: Categorical variables are presented as frequencies (percentages). All the participants with repeated events were included in this study.

**S3 Table.** The impact of Variables on Vascular Events in the Absence of Interaction Note: The generalized linear mixed-effects model (GLMM) was used to evaluate the impact of variables on the vascular when this event changed over time and by participant ID. Statistical significance was set at p < 0.05.

**S4 Table.** The Impact on Vascular Events Over Time in Large Vessels vs. Small & Intermediate Vessels Note: The generalized linear mixed-effects model (GLMM) was used to evaluate the impact of variables on the vascular when this event changed over time and by participant ID. Statistical significance was set at p < 0.05.

**S1 Fig.** The percentage of the first 1-manifestation and first 2-manifestation by year

**S2 Fig.** Yearly Event Percentage Distribution

**S3 Fig.** The Co-occurrence of the First, Second, and Third Events with Vascular Events

